# DAYTIME SLEEPINESS COMPROMISES SPECIFIC PERFORMANCE IN BRAZILIAN JIU-JITSU ATHLETES, WHILE NAPPING IMPROVES SPECIFIC ANAEROBIC CAPACITY: A RANDOMIZED CROSSOVER CLINICAL TRIAL

**DOI:** 10.64898/2026.07.27.26359015

**Authors:** Flávio Aurélio Fernandes Soares, Victor Silveira Coswig, Mario Antônio de Moura Simim, Francisco Adelvane de Paula, Aldo Ângelo Moreira Lima

## Abstract

**Introduction:** Daytime sleepiness (DS) can compromise athletic performance; however, its influence on Brazilian Jiu-Jitsu (BJJ) athletes remains poorly understood. Daytime naps have been suggested as a strategy to alleviate the effects of DS; however, their impact on physical and specific performance remains inconsistent. Thus, this study investigated the influence of different levels of daytime sleepiness on the physical and specific performance of BJJ athletes and verified the effects of daytime napping on these outcomes.

**Methods:** This investigation employed a randomized, controlled, crossover clinical trial design involving 19 amateur Brazilian Jiu-Jitsu (BJJ) athletes who exhibited excessive daytime sleepiness. Participants performed two experimental conditions, with a 30-minute nap (N30) and without a nap (NN), in a randomized order. Vertical jump (CMJ and SJ), anaerobic power (Wingate ), kimono grip strength (KGST), and specific performance were evaluated using the Jiu-Jitsu Anaerobic Performance Test (JJAPT). Data were analyzed using mixed linear models.

**Results:** Daytime sleepiness significantly influenced all parameters of specific performance (JJAPT), with worse performance observed in athletes classified as having abnormal sleepiness. The opportunity to nap increased the total number of repetitions in the JJAPT of four sets (≈20 repetitions; *p* < 0.001), without consistent changes in physical performance tests. An interaction between napping and sleepiness was observed in relation to isometric endurance strength, indicating that napping may provide greater benefits to athletes experiencing abnormal sleepiness.

**Conclusion:** High levels of daytime sleepiness are associated with reduced specific performance in BJJ athletes. Daytime naps are a simple and potentially effective strategy to mitigate these effects and improve specific performance, reinforcing the importance of monitoring sleepiness and individualizing recovery strategies.

## INTRODUCTION

Sleep is an important component in maintaining athletic performance. The physiological processes triggered during sleep contribute to the recovery of athletes ^1,2^. In this sense, due to the high demands of training and competition, it has been recommended that athletes obtain additional sleep time (9 to 10 hours) ^1^compared to healthy adults (7 to 9 hours) ^3^. However, evidence suggests that athletes have insufficient sleep time (± 7.2 hours) ^4^, which is associated with impaired performance ^5^.

In addition to sleep duration, the ability to maintain alertness during wakefulness is considered an important dimension of sleep assessment ^6^. Elevated levels of daytime sleepiness may reflect a non-restorative night’s sleep ^7,8^. Studies on daytime sleepiness have sought to understand its effects on mental health and cognitive performance ^9,10^. In contrast, the effects of sleepiness on athletes’ physical performance remain poorly understood.

On the other hand, daytime naps have been identified as an effective alternative to minimize the negative effects of irregular sleep and daytime sleepiness levels ^11^. A systematic review found that napping consistently reduced daytime sleepiness and increased alertness in athletes ^12^. The authors suggested that napping may provide greater recovery benefits for this population ^12^.

Although there has been a significant increase in the number of studies on sleep and sports in recent years ^13^, the evidence still focuses predominantly on team sports ^14^. Specifically, in combat sports athletes, such as judo, mixed martial arts, taekwondo, karate, and jiu-jitsu, less than 10% of a sample of 2841 participants from 75 studies included in a review addressed this population.

In judo athletes, for example, a four-hour nighttime sleep restriction promoted increased levels of daytime sleepiness and a reduction in anaerobic power in a sprint test ^15^. Conversely, napping attenuates daytime sleepiness and improved anaerobic performance ^15^. Similarly, karate athletes showed reduced levels of daytime sleepiness and improved physical performance after a napping intervention. However, these effects were not observed in a sport-specific performance test ^16^.

Despite these findings, daytime sleepiness has been predominantly used as a complementary measure to assess sleep the night before the intervention, and its influence on athletic performance has been little explored. In this context, the assessment of daytime sleepiness in athletes using subjective instruments, such as the Epworth Sleepiness Scale (ESS) ^17^, can assist coaches and trainers in understanding the athletes’ sleep patterns and their possible influence, in conjunction with daytime napping, on athletic performance. However, studies investigating this relationship are scarce, particularly in Brazilian Jiu-Jitsu (BJJ) athletes. Therefore, this study aimed to investigate the effects of different levels of daytime sleepiness on the physical and specific performance of Brazilian Jiu-Jitsu (BJJ) athletes and to verify the effects of daytime napping on these outcomes. We hypothesized that high levels of daytime sleepiness are associated with reduced physical and specific performance in Brazilian Jiu-Jitsu (BJJ) athletes and that taking a daytime nap mitigates these effects, promoting improved performance.

## MATERIALS AND METHODS

### Participants

Athletes were recruited based on the following inclusion criteria: **i.** attending a minimum of three training sessions per week; **ii.** holding a minimum rank of blue belt. Participants were included if they had participated in at least one official competition of the Brazilian Jiu-Jitsu Confederation (CBJJ). Before the start of the assessments, the participants completed an electronic Google Forms form (Google LLC, California, USA), declaring compliance with the prerequisites: (i) not having performed moderate-or high-intensity physical exercise in the last 24 h and ( ii ) not having ingested stimulants (coffee, energy drinks) or alcoholic beverages in the last 24 h. The exclusion criteria were as follows: **i.** use of sleep medication, **ii.** normal classification for daytime sleepiness assessment (Epworth Sleepiness Scale [ESS]). musculoskeletal injuries that prevented them from performing the physical tests. In total, 19 amateur BJJ athletes (16 men and 3 women) with the following belt levels: blue belt (n = 5), purple belt (n = 6), brown belt (n = 2), and black belt (n = 6) agreed to participate in the study. The study was approved by the ethics committee of the Federal University of Ceará (UFC) under number 6.082.118 (CAAE: 67616923.7.0000.5054).

### Study Design

This was an experimental, randomized, controlled, crossover clinical trial consisting of a control condition (no nap) and an experimental condition (nap). After familiarization with the tests and questionnaires, individuals were randomized to determine the order of conditions: with nap (N30) and without nap (NN) conditions. Before the start of the assessments, participants completed an electronic *Google Forms*® *form* (Google LLC, California, USA), declaring compliance with the prerequisites: **(i)** not having performed moderate or high-intensity physical exercise in the last 24 hours and **(ii )** not having ingested stimulants (coffee, energy drinks) or alcoholic beverages in the last 24 hours.

The intervention consisted of a 30-minute nap (N30) in a private, air-conditioned (22°C) environment with low noise and light levels. Individuals remained alone in the room without access to electronic devices (smartphones*, smartwatches*, or similar devices). The protocol was scheduled to begin at 11:30 a.m., preceding a typical training session (12:00 p.m.). Although this time does not coincide with the circadian sleep propensity (1:00 pm to 3:00 pm), the choice was based on two main aspects: **(i)** the need to adapt the intervention to the athletes’ routine and **(ii )** the possibility of inducing a positive acute effect on recovery before the start of the training session.

The researcher monitored the total intervention time. At the end of the nap and before the start of the tests, the participants declared on a registration form whether they had managed to nap. An additional 30 min was granted to minimize the effects of sleep inertia. For the non-nap period (NN), the participants remained awake for 60 min in another environment, performing passive activities (reading and conversations) under the supervision of a member of the research team. Throughout the study period, the athletes were instructed to maintain an isocaloric diet and usual sleep patterns. Female athletes in the late luteal or early follicular phase (days of menstrual bleeding) were scheduled for a new evaluation due to sleep disturbances reported during this period ^18^.

The assessments were performed in the following order: (1) vertical jump test, (2) anaerobic power assessment, (3) strength endurance assessment, and (4) Jiu-Jitsu anaerobic performance test (JJAPT). Before the start of the assessments, stretching and warm-up exercises were performed (10 min of cycling at a light load, self-selected by the athlete). A 5-minute interval was granted between each test to minimize the effects of residual fatigue and promote a better recovery state. **Figure 1** illustrates the study design. All assessments were performed at the Biomechanics Laboratory of the Institute of Physical Education and Sports of the Federal University of Ceará (IEFES/UFC) from Tuesday to Friday between 11:00 AM and 2:00 PM during August and November 2024.

**Figure 1.**
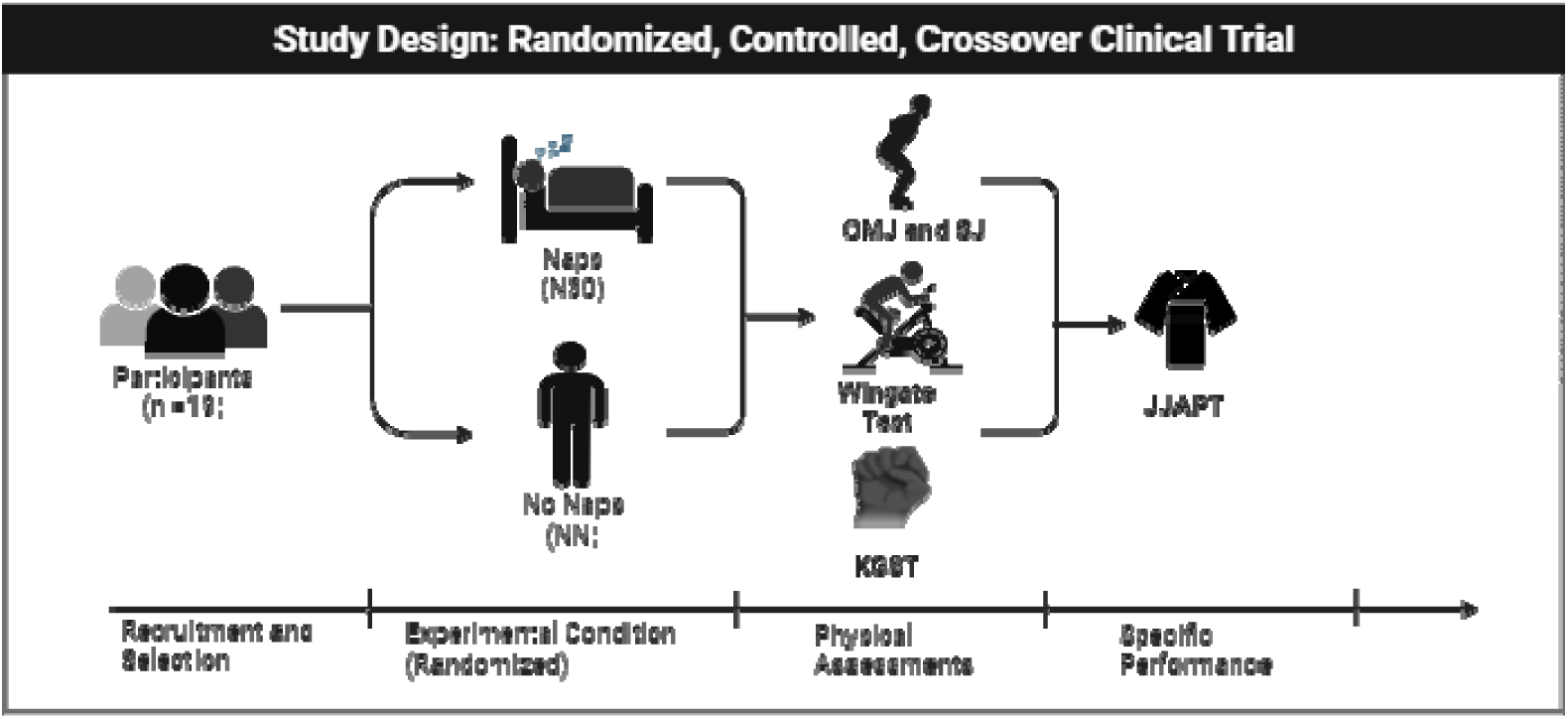
Data collection stage of the study after the familiarization phase of the tests. Nineteen jiu-jitsu athletes (n = 19) were randomized into two experimental conditions in a crossover design: no nap (NN) and 30-minute nap (N30). Subsequently, they underwent physical performance assessments: countermovement vertical jump (CMJ), vertical jump (SJ), anaerobic power ( Wingate Test), kimono grip strength endurance (KGST), and specific performance (Jiu-Jitsu Physical Fitness Test [JJAPT]).

### Instruments

#### Anthropometric Assessment and Training and Competition History

Body mass (kg) and height (m) were measured using a digital scale (200 kg capacity and 100 g increments) with an attached stadiometer (range 115 to 210 cm) - Sanny®. The athletes were instructed to wear light, appropriate clothing for physical exercise and to remove any objects (cell phone, wallet) or accessories (earrings, rings, necklaces). Subsequently, the athletes answered a questionnaire developed by the researchers with information on session duration (in minutes) and training frequency (days of the week) to calculate the weekly training volume, which was the product of training frequency and duration (in minutes).

#### Assessment of Daytime Sleepiness

Daytime sleepiness was assessed using the Epworth Sleepiness Scale. (ESS) ^17^. The ESS consists of eight questions with different contexts (e.g., sitting and reading, watching television, or in public environments) that measure an individual’s subjective probability of falling asleep. For each question, alternatives are proposed – from “would never fall asleep (0)” to “high chance of falling asleep (3)” – whose scores, when added together (ranging from 0 to 24), indicate the individual’s level of daytime sleepiness. The ESS is a reliable instrument for assessing sleepiness and has been translated into Portuguese ^19^.

Although individuals with scores above 10 points (< 10) are classified as having excessive daytime sleepiness, for this study, we opted to use sleepiness subcategories based on the median scores observed in the sample, as recommended in clinical practice.^20^ and previous studies ^21^. For the analyses in the present study, participants who presented an ESS > 10 were classified as having moderate sleepiness (11–15 points) and abnormal sleepiness (16–24 points). This procedure is justified by the ESS being an instrument directly related to the propensity to fall asleep, such that higher scores reflect greater intensity of daytime sleepiness.

#### Vertical Jump Test (CMJ & SJ)

Countermovement jump (CMJ) and squat jump (SJ) tests were performed to assess neuromuscular performance and training readiness. The test protocol consisted of performing three jumps, followed by a 5-second interval between each attempt, to measure the height (in centimeters) of the CMJ and SJ using a contact mat (Jump System Pro; Cefise® - Nova Odessa, São Paulo, Brazil). For the CMJ, the athletes stood on the mat with their knees extended, torso erect, and hands fixed at the hip level. At the evaluator’s signal, the athletes performed flexion followed by extension of the knees as quickly as possible, aiming to reach the highest possible height. For the SJ, athletes started with their knees bent (approximately 90°) and hands on their hips, without any countermovement. The peak and average jump height values were analyzed.

#### 30-Second Wingate test (W30s)

The Wingate test was used to assess maximum anaerobic power and anaerobic capacity using a cycle ergometer (Biotec 2100; Cefise® - Nova Odessa, São Paulo - Brazil). A load relative to the athlete’s body mass (0.075 kg.kg_⁻_¹) was added to the bicycle ^22^. The athletes were instructed to pedal as fast as possible for 30 s in a single attempt. Verbal encouragement was provided to the participants as a form of motivation. The results for maximum power (Pmax), average power (Pmed), minimum power (Pmin), and fatigue index (FI) were calculated using the Ergometric 6.0 softwares. For analysis, only measurements relative to body mass (w.kg_⁻_ ¹) were considered.

#### Kimono Grip Strength Test (KGST)

The isometric resistance force (Maximal *Static) Lift* (MSL) and dynamics ( *Maximum Number of Repetition* – MNR) was assessed using the Kimono *Grip Strength Test* (KGST) as proposed by Franchini et al., (2004) and validated by [reference needed] Silva et al., (2012). For the test, a kimono was placed around a bar suspended at a height of approximately 2 m. In both tests (MSL and MNR), the athletes started from the same position: holding the kimono with their hands and elbow flexion. For the MSL, athletes had to remain suspended statically for as long as possible (in seconds). Only male athletes were evaluated for MNR. The athletes performed the maximum possible number of repetitions, consisting of full extension and flexion of the elbows. Only one attempt was allowed for each test. The KGST can differentiate elite and non-elite athletes ^24^and has shown a high correlation with isokinetic tests [reference needed] ^25^.

#### Specific Anaerobic Performance Test (JJAPT)

The Jiu-Jitsu Anaerobic Test (JJAPT) was developed Villar et al., (2018) and validated by [author’s name (Silva et al., (2019)]. It involves performing the “butterfly” technique for the maximum number of repetitions during 4 or 5 sets of 1 min, with 45 seconds of rest. The athlete should remain lying on their back with their knees bent (approximately 45°) and feet in a “hook” position between their partner’s legs (butterfly guard). The partner – with a similar body mass – will be positioned on their knees, with their spine straight and arms outstretched, adopting a passive posture during the execution of the evaluated movement. The test began with hip and trunk flexion movements (until remaining in a seated position), wrapping the arms around the partner’s trunk. The athlete then moves backward, lifting their partner with the aid of the butterfly guard until the head passes the guard line. Finally, the entire movement is undone, leaving the partner on the mat, and returning to the starting position. For the analysis, protocols with four and five sets were considered for the mean (JJAPTmean), peak (JJAPTpeak), and sum (JJAPTsum) repetition parameters.

### Statistical Analyses

A descriptive analysis was performed to characterize the sample according to the nature of the variables. Subsequently, a linear mixed model (LMM) was used to verify the fixed (main effect and interaction) and random effects on physical and specific performance variables. For the LMM, the normality assumptions of the model residuals were verified using the Shapiro-Wilk test (p < 0.05) and visual inspection of the QQ plot. The homogeneity of the variance of the residuals was evaluated using a residual plot and fitted values. The dependence between repeated measures was considered by including participants as a random effect in the model.

Two models were proposed. Model 1 was fitted with fixed effects considering the sequence of the experimental condition (NN→ N30 or N30 → NN), the period of the condition (1st or 2nd session), the sex of the participants (male and female), and the condition without nap (NN) and with nap (N30) on the physical performance variables. For all proposed models, individuals were considered random intercepts according to the following equation:

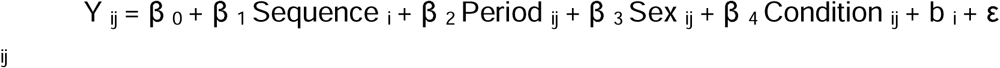

where

- Y _ij:_ dependent variable (physical performance)
- â _0:_ general intercept
- â _1:_ sequence (NN → N30 or N30 → NN)
- â _2:_ period (1st or 2nd session)
- â _3:_ sex (male and female)
- â _4:_ condition without nap (NN) and with nap (N30)
- b _i:_ random effect of the individual (interindividual variability)
- å _ij:_ residual error

Model 2 was fitted with the fixed effect of mean and abnormal sleepiness and the no-nap (NN) and nap (N30) conditions. The model fit with the interaction effect was defined as the sleepiness level and experimental condition (sleepiness*experimental condition). Individuals were considered random effects.

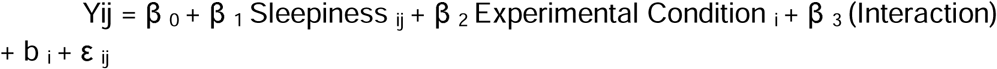

where

- Y _ij:_ dependent variable (physical performance)
- â _0:_ general intercept
- â _1:_ moderate and abnormal sleepiness
- â _2:_ condition without nap (NN) and with nap (N30)
- â _3:_ interaction between drowsiness _and_ experimental condition
- b _i:_ random effect of the individual (interindividual variability)
- å _ij:_ residual error

The values were expressed as estimated marginal means (EMM), with the respective standard error (SE), 95% confidence interval (95% CI), and statistical probability value (p-value). All analyses were performed using JASP Team® software (2025; version 0.95.3) for Windows®, adopting a significance level of 5% (p < 0.05).

## RESULTS

### Sample Characterization and Performance Evaluation between Experimental Conditions (NN and N30) and Daytime Sleepiness

The sample consisted of 19 individuals (n = 19) with an age of 28.2 ± 6.8 years, body mass of 76.7 ± 14.1 kg, and height of 1.70 ± 0.1 m. The individuals had a training experience of 8.7 ± 8.5 years, with a weekly training volume of 258.9 ± 78.8 min. Sixteen participants (84.2%) had competed in state competitions, and three (15.8%) athletes stated that they had competed in national competitions. Twelve (n = 12) athletes presented with abnormal sleepiness, and seven (n = 7) presented with moderate sleepiness.

### Analysis with Model Adjustment 1 - Sequence of the Experimental Condition: Period of the Condition, Sex of the Participants, and Experimental Condition

The linear mixed model (LMM) was fitted for the sequence variables of the experimental conditions (NN → N30 or N30 → NN), condition period (1st or 2nd session), sex (male and female), and experimental condition to examine the fixed and interaction effects on physical and specific performance parameters. Subjects were treated as random effects because of their reliance on repeated measures. The model demonstrated a statistically significant influence of sex on physical performance tests, with the exception of the minimum anaerobic power (P. Min). and isometric endurance strength (MSL) were measured. For the other outcomes, the male participants showed superior performance compared to the female participants.

For the specific performance variables, the LMM showed a significant effect only of the experimental condition (NN vs. N30) on the sum of repetitions in the specific anaerobic jiu-jitsu test in four sets (JJAPTsum4). **Table 1** presents the results of the statistical analyses of fixed effects for the physical and specific performance variables with the estimates of the fixed effects coefficients (β), the respective standard errors (SE), and *p-values* obtained for the statistically significant variables.

**Table 1.** Results of the linear mixed model for physical and specific performance variables for Model 1. F (p-value). Coefficients of fixed effects (β) and estimated marginal means ( EMMs ), respectively, of the effect of sex on physical performance indicators and of the experimental condition on specific performance indicators.

| PHYSICAL PERFORMANCE | SEQUENCE | PERIOD | SEX | CONDITION | $\beta$ (SE) | p value | FAMELE | | MALE | |
| --- | --- | --- | --- | --- | --- | --- | --- | --- | --- | --- |
|  |  |  |  |  |  |  | EMM (SE) | 95% CI | EMM (SE) | 95% CI |
| CMJmean (cm) | 0.03 (0.85) | 0.95 (0.34) | **5.63 (0.03)** | 2.80 (0.11) | 9.63 (4.05) | 0.03 | 23.44 (3.67) | 16.25 to 30.64 | 33.08 (1.61) | 29.91 to 36.25 |
| CMJpico (cm) | 0.15 (0.70) | 0.05 (0.82) | **5.53 (0.03)** | 2.26 (0.15) | 9.59 (4.0) | 0.03 | 24.79 (3.68) | 17.56 to 32.02 | 34.38 (1.62) | 31.20 to 37.57 |
| SJmean (cm) | 0.20 (0.65) | 0.02 (0.88) | **6.94 (0.01)** | 0.38 (0.54) | 9.30 (3.53) | 0.01 | 22.95 (3.19) | 16.69 to 29.21 | 32.25 (1.40) | 29.49 to 35.01 |
| SJpico (cm) | 0.20 (0.65) | 0.00 (0.96) | **7.11 (0.01)** | 0.68 (0.42) | 9.37 (3.51) | 0.01 | 23.95 (3.17) | 17.72 to 30.18 | 33.33 (1.40) | 30.58 to 36.07 |
| P. Max (W.kg) | 0.63 (0.43) | 0.51 (0.48) | **8.7 (0.00)** | 3.78 (0.06) | 2.35 (0.79) | 0.00 | 9.59 (0.72) | 8.17 to 11.01 | 11.95 (0.31) | 11.32 to 12.57 |
| P. Med (W.kg) | 0.14 (0.70) | 0.02 (0.87) | **8.94 (0.00)** | 1.11 (0.3) | 1.41 (0.47) | 0.00 | 7.29 (0.42) | 6.45 to 8.12 | 8.70 (0.18) | 8.33 to 9.07 |
| P. Min (W.kg) | 0.68 (0.80) | 0.26 (0.61) | 0.16 (0.69) | 1.08 (0.30) | X | X | X | X | X | X |
| IF (%) | 1.15 (0.29) | 0.97 (0.33) | **6.58 (0.01)** | 2.39 (0.13) | 14.30 (5.57) | 0.01 | 41.24 (5.04) | 31.35 to 51.12 | 55.54 (2.22) | 51.18 to 59.90 |
| MNR (rep/kg) | 0.79 (0.38) | 0.60 (0.45) | NA | 0.46 (0.50) | NA | NA | NA |  | NA |  |
| MSL (sec /kg) | 0.09 (0.76) | 1.46 (0.24) | 0.51 (0.48) | 1.57 (0.22) | X | X | X | X | X | X |
| SPECIFIC PERFORMANCE | SEQUENCE | PERIOD | SEX | CONDITION | $\beta$ (SE) | p value | NO NAP (NN) | | NAP (N30) | |
|  |  |  |  |  |  |  | EMM (SE) | 95% CI | EMM (SE) | 95% CI |
| APT mean4 (rept) | 1.33 (0.26) | 0.50 (0.48) | 0.53 (0.47) | 1.87 (0.18) |  |  |  |  |  |  |
| APTmean5 (rept) | 1.42 (0.24) | 0.55 (0.46) | 0.40 (0.53) | 2.42 (0.13) | X | X | X | X | X | X |
| APTpeak4 (rept) | 0.19 (0.66) | 0.11 (0.73) | 0.00 (0.94) | 0.95 (0.34) | X | X | X | X | X | X |
| APTpeak5 (rept) | 0.26 (0.61) | 0.00 (0.95) | 0.05 (0.82) | 2.49 (0.13) | X | X | X | X | X | X |
| APTsum4 (rept) | 1.06 (0.31) | 0.69 (0.41) | 0.64 (0.43) | **118.23 (0.00)** | -20.09 (1.84) | 0.00 | 74.98 (3.34) | 68.42 to 81.53 | 95.06 (3.34) | 88.51 to 101.6 |
| APTsum5 (rept) | 1.42 (0.24) | 0.55 (0.46) | 0.40 (0.53) | 2.42 (0.13) | X | X | X | X | X | X |

**SEQUENCE:** NN to N30 or N30 to NN. **PERIOD:** 1st session or 2nd session. **GENDER:** Male and Female. **CONDITION:** Without nap (NN) and with nap (N30). **F (p-value):** Result of statistical analysis and significance values. **CMJ mean (cm):** Average countermovement vertical jump (centimeters). **CMJpeak (cm):** Maximum height of the countermovement vertical jump (in centimeters). **SJmean (cm):** Height Average vertical jump height (cm). **SJpeak:** Maximum vertical jump height (cm). **P. Max (W.Kg):** Maximum power (watts per kilogram); **P. Med (W.Kg):** Average power (watts per kilogram); **P. Min (W.Kg · kg):** Minimum power (watts per kilogram). **IF:** Fatigue index (percentage); **MNR (rep/kg):** Relative dynamic resistance strength (number of repetitions per kg). **MSL (sec/kg):** Relative isometric resistance strength (s/kg). **JJAPTmean4 (rept):** Average number of repetitions in the

**Figure 2** shows the dispersion of individuals from the intercept and the marginal means (standard error) of the JJAPTsum4 results between the conditions (NN and N30). A greater total number of repetitions was observed in the nap condition (N30 – blue circle) with estimated marginal means of 95 repetitions (95% CI: 88–101) compared to the no-nap condition (NN) with 74 repetitions (95% CI: 68–81), indicating an average difference of approximately 20 repetitions between the conditions.

**Figure 2.**
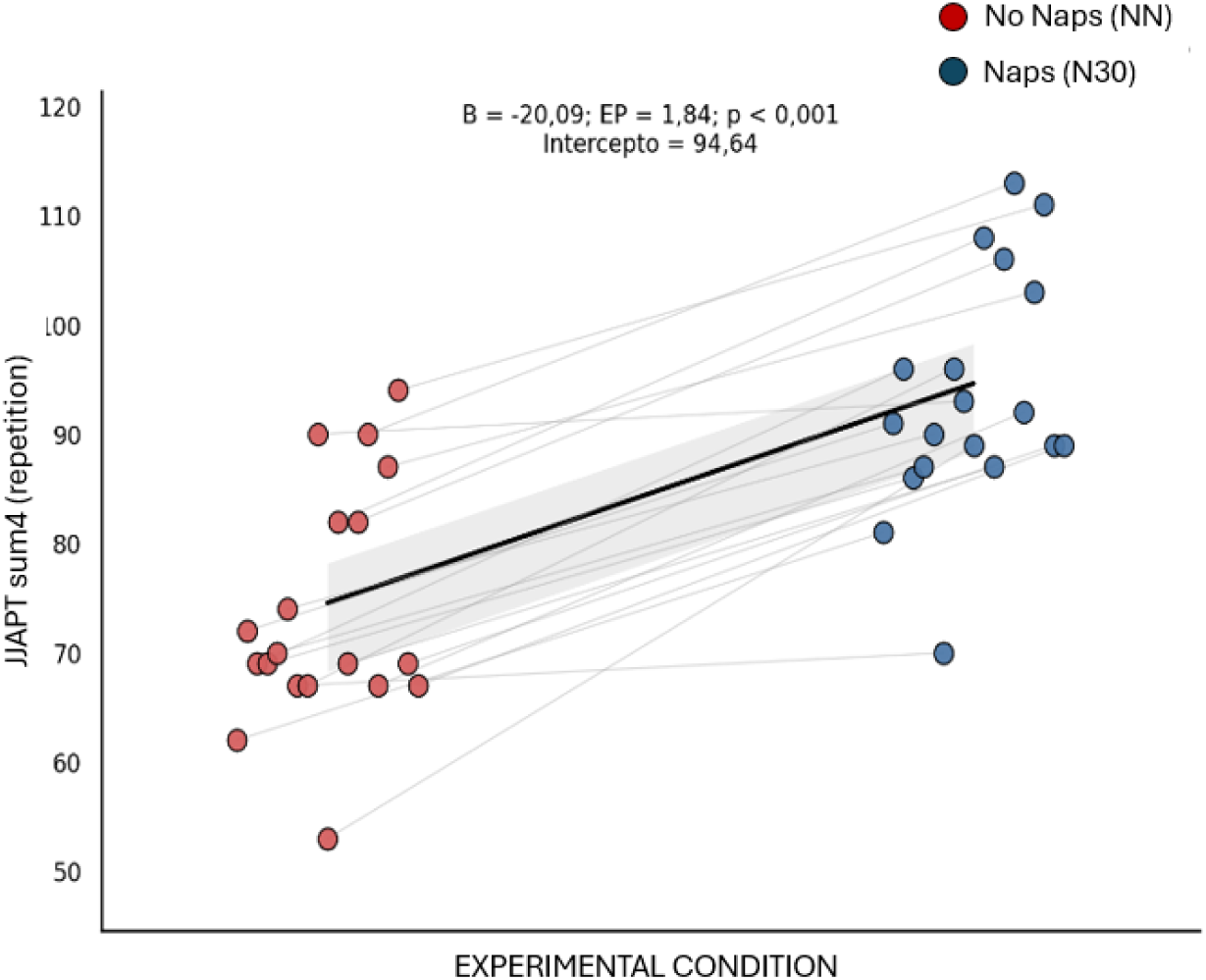
Dispersion of individuals among the experimental conditions indicating superior performance in the nap condition (blue circle) compared to the no-nap condition (red circle). **Black line:** intercept of the model (94.64 repetitions) with the nap condition (N30) as the reference standard, representing the estimated average value of the specific performance test (JJAPT sum4).

### Analysis with Model Adjustment 2 - Experimental Condition and Daytime Sleepiness

The model was fitted with the experimental condition (no nap (NN) and nap (N30] ), the level of daytime sleepiness (abnormal sleepiness and average sleepiness), and the interaction (experimental condition × daytime sleepiness) to assess the effect on physical and specific performance. Individuals were included as random effects. The results did not indicate a main effect on physical performance variables.

For the specific performance variables, the effect of the experimental condition (NN and N30) remained for the total number of repetitions for four series (JJAPT sum4) with F _(1,17)_ = 105.6 (p < 0.001) with higher values in the nap condition (N30) with 94.93 repetitions (95% CI 90.44 to 99.43) compared to the no-nap condition (NN) with 75.12 repetitions (95% CI 70.62 to 79.61) with a mean difference of approximately 20 repetitions.

Additionally, the model indicated a statistically significant effect of daytime sleepiness level on all specific performance parameters: mean repetitions (JJAPT mean), highest repetition (JJAPT peak), and total number of repetitions (JJAPT sum) for protocols with four and five sets. The results demonstrated that individuals with abnormal sleepiness performed worse than those with average sleepiness, suggesting that the level of sleepiness can influence the performance of jiu-jitsu athletes. **Figure 3** shows the dispersion of individuals from the intercept and the marginal means (standard error) of the specific performance test results (JJAPT) according to the sleepiness level classification.

**Figure 3.**
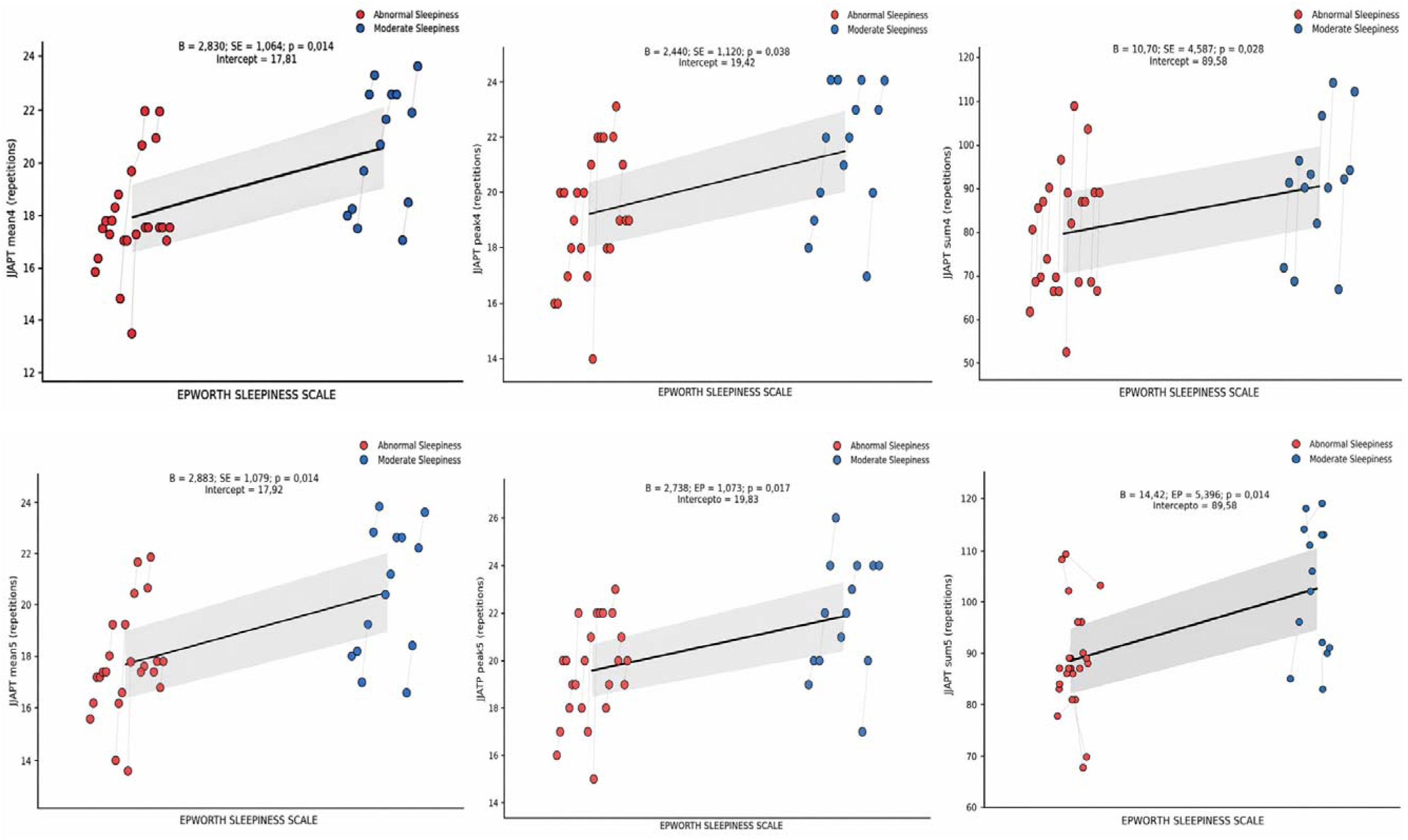
Dispersion of individuals among the experimental conditions with the model adjustment based on the assessment of sleepiness levels: NN, no nap (red circle); N30, with nap (blue circle). **Black line:** model intercept. Individuals classified as having average sleepiness performed better than those with abnormal sleepiness on all parameters of specific performance (JJAPT mean, JJAPT peak, and JJAPT sum).

Finally, the model showed an interaction effect (experimental condition * daytime sleepiness) for isometric resistance strength (MSL) with F _(1, 16)_ = 4.56 (p = 0.048). The results indicated that the effect of the condition varied according to the level of sleepiness. In the no-nap condition (NN), individuals exhibiting moderate sleepiness demonstrated superior performance compared to those with abnormal sleepiness, with a difference of approximately 0.20 seconds. However, in the nap condition (N30), this difference between the groups (abnormal and moderate sleepiness) was reduced, with similar values. Furthermore, individuals exhibiting abnormal sleepiness in the N30 condition demonstrated an improvement of 0.128 s in performance compared to those in the NN condition. In other words, a 30-minute nap appears to be associated with better performance in isometric strength, especially in individuals with abnormal sleepiness. **Figure 4** presents the marginal means (standard error) of the interaction between the experimental conditions (NN and N30) and the level of sleepiness (abnormal and normal).

**Figure 4.**
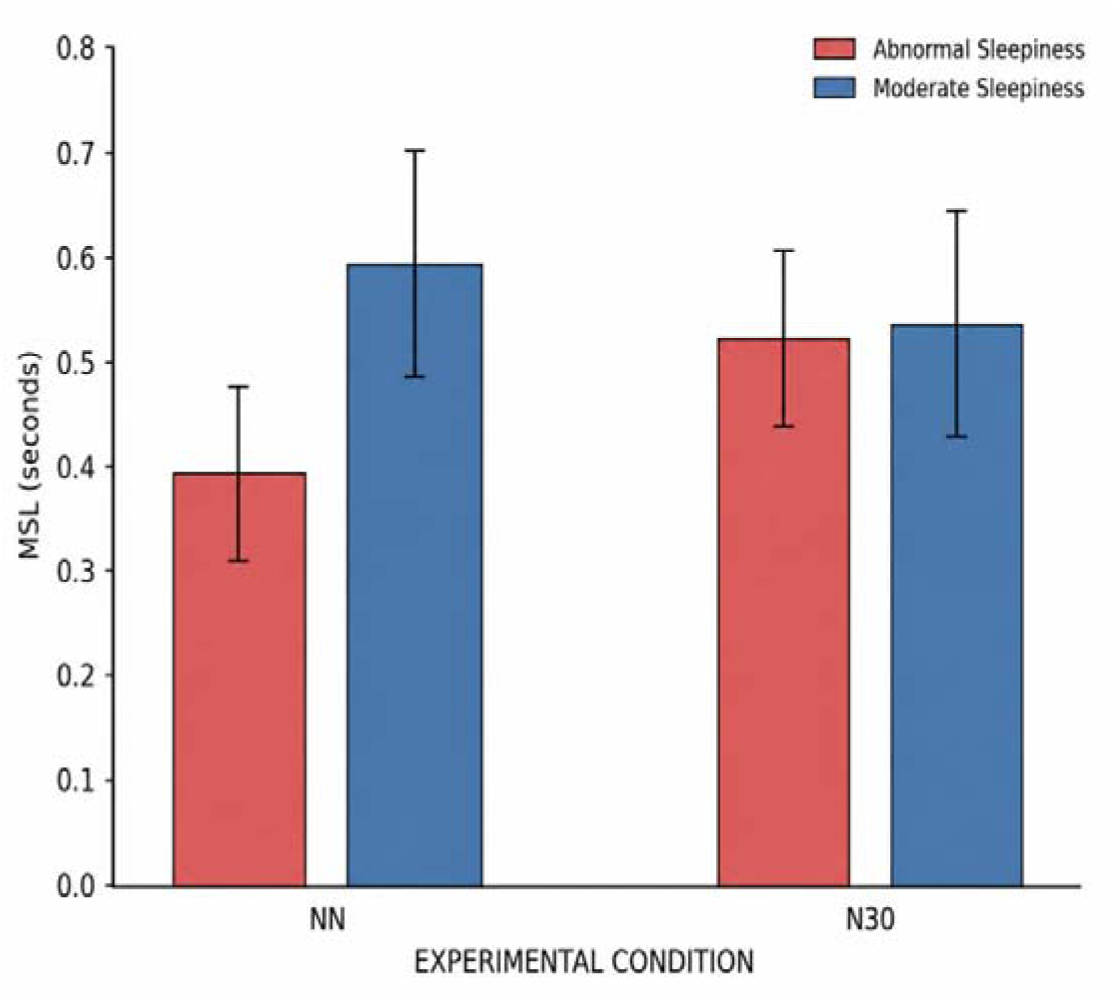
Interaction between experimental conditions (NN and N30) and daytime sleepiness levels (abnormal and average) in the isometric resistance strength test (MSL). Individuals with abnormal sleepiness (in red) performed better after a nap, suggesting a positive effect of the N30 condition on isometric performance. In contrast, individuals with average sleepiness (in blue) showed superior performance in the no-nap condition (NN), while in the (N30) condition, the averages between the sleepiness groups were similar. The bars represent the marginal means estimated by the model, and the error lines indicate the standard errors.

## DISCUSSION

This study aimed to investigate the effect of different levels of daytime sleepiness on the physical and specific performance of Brazilian Jiu-Jitsu (BJJ) athletes and to verify the effect of daytime napping on these outcomes. The results partially corroborated the proposed hypothesis, indicating that daytime sleepiness influences athletes’ specific performance but not their physical performance. Additionally, the effects of napping were observed only for specific performances without promoting consistent changes in physical performance variables.

Daytime sleepiness is defined as the tendency or propensity to fall asleep during the day ^7^. Individuals with high levels of sleepiness have difficulty staying awake and maintaining alertness throughout the day ^7^. The homeostatic component of sleep has been identified as a strong influence on sleepiness levels ^28^. This component is associated with low quantity and/or quality of sleep or early awakening ^7,29^, increasing wakefulness time.

Prolonged wakefulness increases the body’s energy demand. Adenosine triphosphate (ATP) is the main molecule responsible for energy transfer during cellular metabolism ^30^. The hydrolysis of ATP results in an increase in extracellular adenosine concentration, which is recognized as a crucial endogenous marker of homeostatic sleep pressure 31. Adenosine accumulation is associated with an increased subjective perception of effort ^32^. Although sleepiness and fatigue are distinct constructs, these conditions have a bidirectional relationship, such that changes in one can affect the other ^33^. In this context, it is plausible to assume that elevated levels of daytime sleepiness may modulate the perception of effort, intensifying the sensation of fatigue and compromising the ability to sustain specific performance in tasks involving repeated high-intensity efforts, such as the specific performance test used in this study (JJAPT).

While the opportunity for a nap did not decrease levels of daytime sleepiness, the findings consistently indicated an effect of the nap on specific performance, as measured by the JJAPT sum 4. In this context, it has been suggested that performance assessments based on sport-specific technical movements may be more sensitive in detecting the effects of sleep-related interventions in athletes than general physical performance tests ^5,34–36^, which is consistent with our findings.

Furthermore, an interesting point in our results was that the nap effect was observed only in the sum of repetitions (JJAPTsum) without concomitant changes in the peak (JJAPTpeak) or average (JJAPTmean) parameters. The total volume of work performed during the evaluation was represented by the sum of repetitions. This measure appears to be more sensitive in detecting discrete changes resulting from sleep-related interventions in specific tasks. However, the absence of a nap effect in a protocol with five sets (JJAPTsum5) may be related to the greater accumulation of fatigue throughout the task and/or the influence of adopting self-regulation strategies for effort and adjusting the execution pace according to the duration of the test.

This study has some limitations that should be considered in the interpretation of the results, such as the absence of physiological measures related to sleep and fatigue mechanisms, the use of subjective instruments for the evaluation of napping, and the absence of sleep monitoring the night before the experimental sessions, which may have influenced daytime sleepiness levels and, consequently, the athletes’ performance. Therefore, future studies should incorporate objective sleep measures, such as actigraphy or polysomnography, and physiological markers that allow for a more precise understanding of the mechanisms involved in the relationship between daytime sleepiness, napping, and athletic performance. From a practical standpoint, the results suggest that coaches and trainers should monitor the daytime sleepiness levels of their athletes, as this condition can compromise their performance. Furthermore, they should consider incorporating a daytime nap into their training routine as a recovery strategy to mitigate the effects of daytime sleepiness and improve specific performance in Brazilian Jiu-Jitsu athletes.

## CONCLUSION

The findings of this study demonstrate that daytime sleepiness is a factor associated with the specific performance of Brazilian Jiu-Jitsu athletes, since individuals with higher levels of sleepiness showed inferior performance in specific tests of the sport. Furthermore, an opportunity for a daytime nap improved specific performance, especially in the total volume of work performed (JJAPTsum4), although it did not promote consistent changes in indicators of general physical performance. Taken together, these results suggest that interventions aimed at managing daytime sleepiness, such as the strategic inclusion of naps in the training routine, may represent a simple and potentially effective approach to optimize specific performance in Brazilian Jiu-Jitsu athletes. However, the observed effects seem to depend on the outcome assessed and the athlete’s level of sleepiness, reinforcing the importance of individualized sleep monitoring and recovery strategies in the sports context.

## Data Availability

The datasets generated and analyzed during this study are available from the corresponding author upon reasonable request, subject to ethical and institutional requirements.

## ACKNOWLEDGEMENTS

The authors thank the athletes who participated in the study for their availability and collaboration throughout all stages of this research. They also thank the institutions and laboratories involved for the structural and scientific support provided for this study. This manuscript is original, has not been previously published, and is not being considered for publication in any other journal. All the authors have approved the submission. The corresponding author has an active affiliation with the Federal University of Ceará (Brazil) and meets the eligibility criteria of the CAPES “ Read & Publish “ agreement. After acceptance, we will opt for publication in Open Access under the CC BY license.

